# Genetic Evidence for a Causal Link between Intra-Pancreatic Fat Deposition and Pancreatitis: a Mendelian Randomization Study

**DOI:** 10.1101/2024.06.03.24308330

**Authors:** Hajime Yamazaki, Martin Heni, Róbert Wagner, Shunichi Fukuhara, Steven R. Grossman, Sihao Han, Lang Wu, Samantha A. Streicher, Brian Z. Huang

## Abstract

**Introduction:** Recent associative studies have linked intra-pancreatic fat deposition (IPFD) with risk of pancreatitis, but the causal relationship remains unclear.

**Methods:** Utilizing Mendelian randomization, we evaluated the causal association between genetically predicted IPFD and pancreatitis. This approach utilized genetic variants from genome-wide association studies of IPFD (n=25,617), acute pancreatitis (n=6,787 cases/361,641 controls), and chronic pancreatitis (n=3,875 cases/361,641 controls).

**Results:** Genetically predicted IPFD was significantly associated with acute pancreatitis (OR per 1-SD increase: 1.40[95%CI:1.12-1.76], p=0.0032) and chronic pancreatitis (OR:1.64[95%CI:1.13-2.39], p=0.0097).

**Discussion:** Our findings support a causal role of IPFD in pancreatitis, suggesting that reducing IPFD could lower the risk of pancreatitis.

## Introduction

Intra-pancreatic fat deposition (IPFD) represents a state of lipid accumulation within the pancreas, which can be detected by ultrasonography, computed tomography, and magnetic resonance imaging (MRI)(1, 2). Our previous prospective cohort study demonstrated an association between IPFD and pancreatic cancer, and our subsequent Mendelian randomization (MR) analysis provided causal evidence for its pathogenic impact(3). MR analysis is regarded as a natural randomized trial to assess causality since it uses genetic variants, which are randomly assigned at conception and remain unaffected by subsequent acquired diseases or environmental influences. This approach minimizes issues of reverse causation and confounding present in observational studies(3-5).

Of note, a recent prospective cohort study not only verified the relationship between IPFD and pancreatic cancer but also revealed an association with acute pancreatitis (AP), which is one of the leading causes of gastrointestinal-related hospital admissions(6, 7). To further elucidate causality between IPFD and pancreatitis, we conducted a two-sample MR study to evaluate the causal association of IPFD with both AP and chronic pancreatitis (CP).

## Methods

We leveraged eight independent genetic variants associated with MRI-measured IPFD (p<5× 10^−8^) from a genome-wide association study (GWAS) using UK Biobank data (n=25,617 individuals)(8). In this study, the mean age (standard deviation [SD]) was 64 (7.5) years, and 51% of the participants were female. The mean (SD) of IPFD was 10.4 (7.9) %(8). We assessed the association of these IPFD-associated genetic variants with AP and CP using data from the GWAS of AP (6,787 cases and 361,641 controls) and CP (3,875 cases and 361,641 controls) in the FinnGen study(9). Diagnoses of AP and CP were identified using ICD or KELA reimbursement codes (see: https://r10.risteys.finngen.fi/endpoints/K11_ACUTPANC and https://r10.risteys.finngen.fi/endpoints/K11_CHRONPANC). Ethical review for the current analyses was waived as the data utilized were de-identified and sourced from public databases.

For the primary MR analyses, the association of each IPFD-associated genetic variant with AP or CP was weighted by its association with IPFD, and estimates were combined using the inverse variance weighted (IVW) method. To evaluate the strength of the association between each IPFD-associated genetic variant and IPFD, we calculated F-statistics, which values >10 indicating valid genetic variants for MR(4). Cochran’s Q value was calculated to evaluate the heterogeneity among estimates obtained using different genetic variants.

To account for potential pleiotropy (i.e., IPFD-associated genetic variants affecting pancreatitis through pathways other than IPFD), we evaluated whether these IPFD associated genetic variants were also associated with alcohol use or cholelithiasis using published GWAS data(10, 11). None of the eight IPFD-associated genetic variants were associated with alcohol use or cholelithiasis at a GWAS threshold (p < 5×10^−8^). However, using the Bonferroni corrected threshold of p<0.00625 (p<0.05/8 genetic variants), as used in previous MR studies(3, 12), one genetic variant (rs115478735) had a nominal association with cholelithiasis. Therefore, leave-one-out analyses was conducted by excluding each genetic variant from the MR analysis. Furthermore, we conducted other sensitivity analyses, including pleiotropy-robust MR methods (weighted median and MR-Egger) and multivariable MR adjusting for visceral adipose tissue (VAT) volume using GWAS data(8).

Finally, we evaluated the possibility of reverse causation (i.e., pancreatitis causing IPFD rather than IPFD causing pancreatitis) using bidirectional MR. Utilizing genetic variants associated with AP and CP (p<5×10^−8^)(9), we assessed their association with IPFD using corresponding GWAS data(8).

We considered p<0.05 as statistically significant for our MR analyses. Statistical analyses were performed using Stata18, R version-4.3.3, the TwoSampleMR package, and MendelianRandomization package.

## Results

All eight genetic variants were strongly associated with IPFD: mean F-statistics 52 (min 32, max 95). Genetically predicted IPFD was associated with risks for both AP and CP (**Figure 1**). Per-1 SD increase in IPFD, there was a 40% increased odds of AP (odds ratio [OR] 1.40, 95% confidence interval [95%CI]:1.12-1.76, p=0.0032), and 64% increased odds of CP (OR 1.64, 95%CI:1.13-2.39, p=0.0097). Sensitivity analyses including VAT-adjusted multivariable MR were largely consistent with this finding (**Table 1**).

**Table 1.**
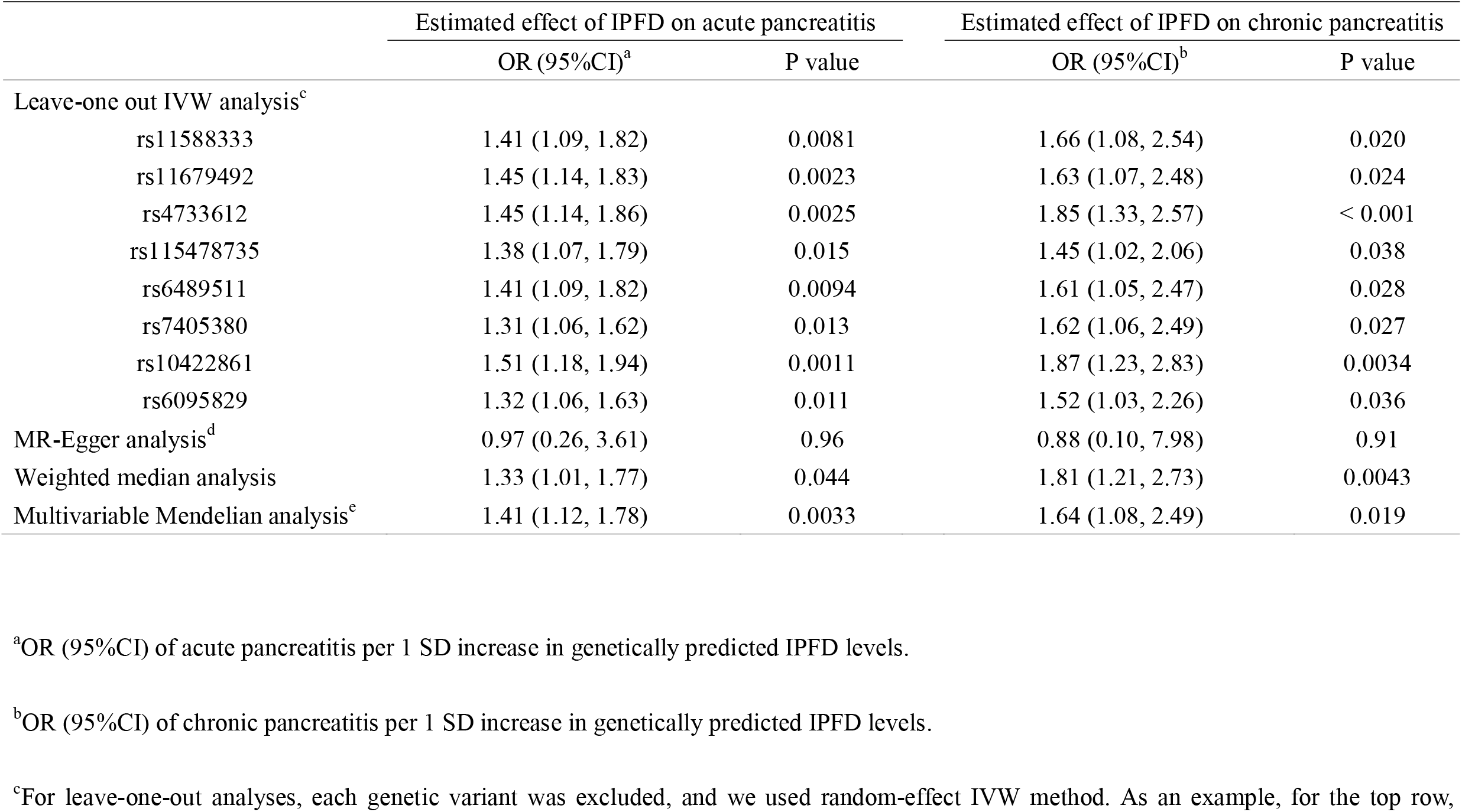
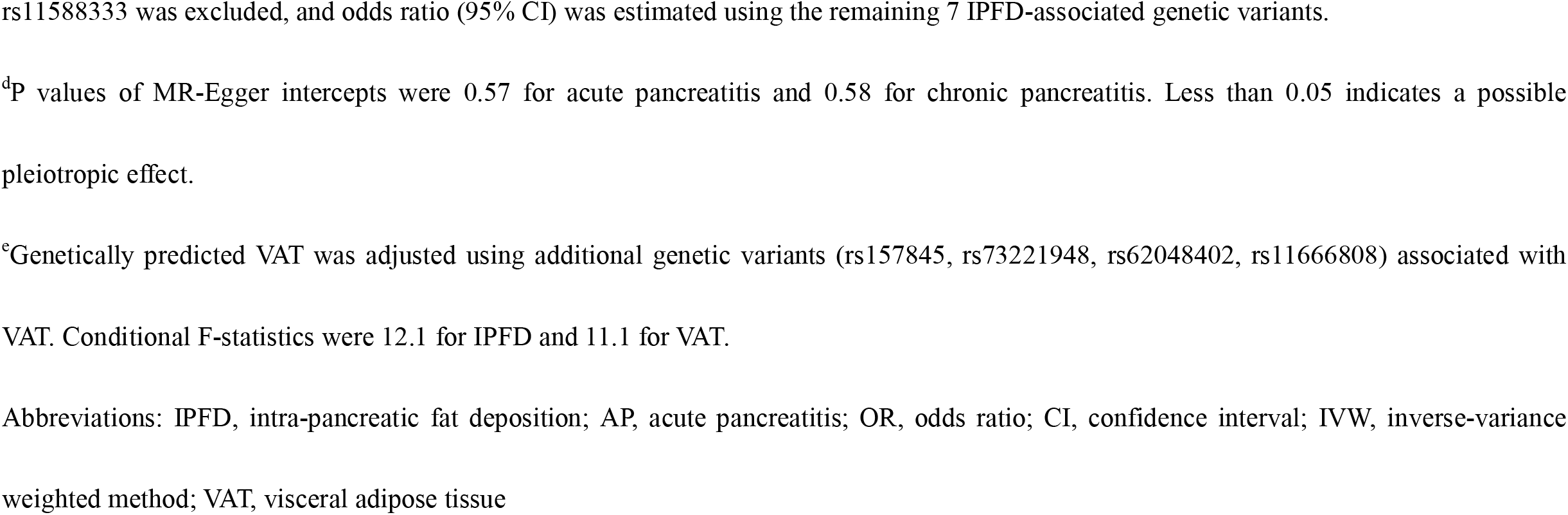
Estimates from Sensitivity Mendelian Randomization analyses for the effect of intra-pancreatic fat deposition on acute and chronic pancreatitis.

|  | Estimated effect of IPFD on acute pancreatitis |  | Estimated effect of IPFD on chronic pancreatitis |  |
| --- | --- | --- | --- | --- |
|  | OR (95%CI)<sup>a</sup> | P value | OR (95%CI)<sup>b</sup> | P value |
| Leave-one out IVW analysis<sup>c</sup> |  |  |  |  |
| rs11588333 | 1.41 (1.09, 1.82) | 0.0081 | 1.66 (1.08, 2.54) | 0.020 |
| rs11679492 | 1.45 (1.14, 1.83) | 0.0023 | 1.63 (1.07, 2.48) | 0.024 |
| rs4733612 | 1.45 (1.14, 1.86) | 0.0025 | 1.85 (1.33, 2.57) | < 0.001 |
| rs115478735 | 1.38 (1.07, 1.79) | 0.015 | 1.45 (1.02, 2.06) | 0.038 |
| rs6489511 | 1.41 (1.09, 1.82) | 0.0094 | 1.61 (1.05, 2.47) | 0.028 |
| rs7405380 | 1.31 (1.06, 1.62) | 0.013 | 1.62 (1.06, 2.49) | 0.027 |
| rs10422861 | 1.51 (1.18, 1.94) | 0.0011 | 1.87 (1.23, 2.83) | 0.0034 |
| rs6095829 | 1.32 (1.06, 1.63) | 0.011 | 1.52 (1.03, 2.26) | 0.036 |
| MR-Egger analysis<sup>d</sup> | 0.97 (0.26, 3.61) | 0.96 | 0.88 (0.10, 7.98) | 0.91 |
| Weighted median analysis | 1.33 (1.01, 1.77) | 0.044 | 1.81 (1.21, 2.73) | 0.0043 |
| Multivariable Mendelian analysis<sup>e</sup> | 1.41 (1.12, 1.78) | 0.0033 | 1.64 (1.08, 2.49) | 0.019 |

<sup>a</sup>OR (95%CI) of acute pancreatitis per 1 SD increase in genetically predicted IPFD levels.
<sup>b</sup>OR (95%CI) of chronic pancreatitis per 1 SD increase in genetically predicted IPFD levels.
<sup>c</sup>For leave-one-out analyses, each genetic variant was excluded, and we used random-effect IVW method. As an example, for the top row,
rs11588333 was excluded, and odds ratio (95% CI) was estimated using the remaining 7 IPFD-associated genetic variants.
<sup>d</sup>P values of MR-Egger intercepts were 0.57 for acute pancreatitis and 0.58 for chronic pancreatitis. Less than 0.05 indicates a possible pleiotropic effect.
<sup>e</sup>Genetically predicted VAT was adjusted using additional genetic variants (rs157845, rs73221948, rs62048402, rs11666808) associated with VAT. Conditional F-statistics were 12.1 for IPFD and 11.1 for VAT.
Abbreviations: IPFD, intra-pancreatic fat deposition; AP, acute pancreatitis; OR, odds ratio; CI, confidence interval; IVW, inverse-variance weighted method; VAT, visceral adipose tissue

**Figure 1.**
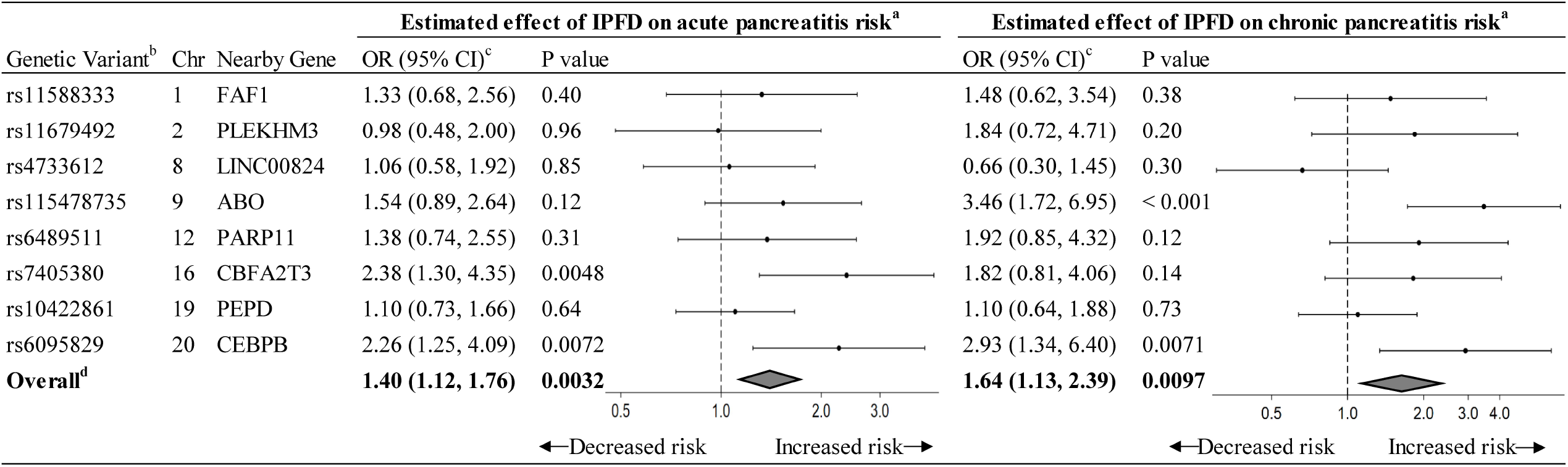
Mendelian randomization estimates for the effect of intra-pancreatic fat deposition on acute and chronic pancreatitis. ^a^Data markers indicate the OR for the association of genetically predicted IPFD with acute pancreatitis or chronic pancreatitis, which was estimated using each genetic variant. Error bars indicate 95% CIs. ^b^Genetic variants associated with IPFD levels. Mean F-statistic was 52. ^c^OR (95%CI) of acute pancreatitis or chronic pancreatitis per 1 SD increase in genetically predicted IPFD levels ^d^Random-effect inverse-variance weighted method was used to obtain overall estimate for the association of genetically predicted IPFD with pancreatitis. Cochran’s Q values were 8.7 for acute pancreatitis and 14.1 for chronic pancreatitis. Abbreviations: IPFD, intra-pancreatic fat deposition; Chr, chromosome; OR, odds ratio; CI, confidence interval; SD, standard deviation

Bidirectional MR did not support reverse causation for either AP (p=0.32) or CP (p=0.15) (**Figure 2**).

**Figure 2.**
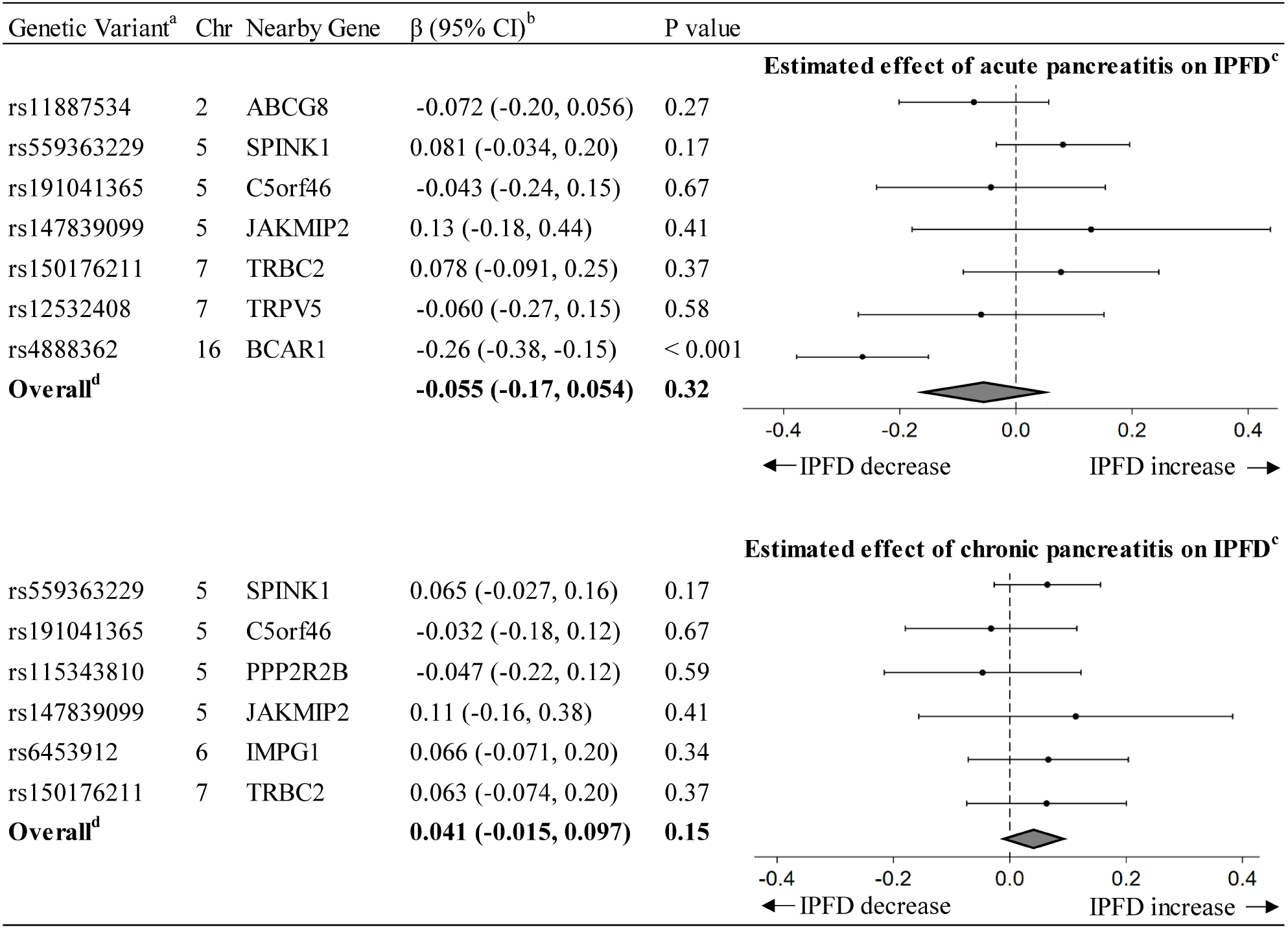
Bidirectional Mendelian randomization estimates for the effects of acute and chronic pancreatitis on intra-pancreatic fat deposition. ^a^Genetic variants associated with acute pancreatitis or chronic pancreatitis. Mean F-statistics were 65.2 for acute pancreatitis and 60.8 for chronic pancreatitis. ^b^β (95%CI) in SD units for the estimated effect of genetically predicted acute pancreatitis or chronic pancreatitis on IPFD. ^c^Data markers indicate the β for the association of genetically predicted acute pancreatitis or chronic pancreatitis with IPFD, which was estimated using each genetic variant. Error bars indicate 95% CIs. ^d^Random-effect inverse-variance weighted method was used to obtain overall estimated effect of genetically predicted acute pancreatitis or chronic pancreatitis on IPFD. Cochran’s Q values were 22.3 for acute pancreatitis and 2.7 for chronic pancreatitis. Abbreviations: IPFD, intra-pancreatic fat deposition; Chr, chromosome; β, β coefficient; CI, confidence interval; SD, standard deviation

## Discussion

Our bidirectional MR study provides evidence for a causal impact of IPFD on both AP and CP. This relationship likely exists independently of general visceral fat accumulation, as indicated by the VAT-adjusted multivariable MR analysis.

Based on the findings from our MR study and corroborative evidence from previous observational studies(1, 6, 13), IPFD is likely a pathophysiological contributor to pancreatitis. In addition to several cross-sectional studies(1), a cohort study with about 10,000 participants undergoing ultrasonography with a 6-year follow-up demonstrated an association between IPFD and CP(13). Furthermore, a recent prospective cohort study involving more than 40,000 participants receiving MRI with a 4.6-year follow-up showed an association between IPFD and AP(6). Potential mechanisms linking IPFD to pancreatitis include lipid overload causing cellular stress responses, lipotoxicity, and aberrant cellular crosstalk, leading to the activation of immune cells and stellate cells(1). Notably, IPFD is a modifiable condition and can be reduced, for example, through dietary interventions(14).

One major strength of this study is the use of bidirectional MR and multivariable MR, which are robust approaches that are less susceptible to reverse causation and confounding(4). Furthermore, we utilized data from the largest available GWAS on IPFD. We also used IPFD data measured with MRI, which has been well validated against histological measurements of IPFD(15).

There are some limitations to our study. The unknown mechanism linking the genetic variants, IPFD, and pancreatitis may involve pleiotropic effects. To minimize the influence of potential pleiotropic effects, we confirmed the robustness of our results through several sensitivity analyses. Moreover, this study was limited to individuals of European ancestry.

In conclusion, IPFD is a novel risk factor that likely plays a causal role in the development of pancreatitis. The evidence from our MR study, combined with previous associative data, supports the possibility that reducing IPFD could lower the risk of pancreatitis.

## Data Availability

GWAS data for IPFD, VAT, alcohol use, and cholelithiasis are accessible through the GWAS Catalog (https://www.ebi.ac.uk/gwas/). GWAS data for pancreatitis are available from FinnGen (https://www.finngen.fi/en).

## Acknowledgements

We want to acknowledge the participants and investigators of the FinnGen study and UK Biobank.

## Author contributions

H.Y. designed the study, collected the data, and wrote the draft. H.Y. and S.H. conducted the analysis. H.Y., M.H., R.W., S.F., S.R.G., S.H., L.W., S.A.S., and B.Z.H. reviewed, made critical revisions, and approved the article before submission. H.Y. is the guarantor of this work and, as such, had full access to all the data in the study and takes responsibility for the integrity of the data and the accuracy of the data analysis.

